# Association Between use of an Adaptive Cardiac Resynchronization Therapy Algorithm and Healthcare Utilization and Cost

**DOI:** 10.1101/2023.12.08.23299751

**Authors:** Michael R. Gold, Jiani Zhou, Lucas Higuera, David P. Lanctin, Eugene S. Chung

**Affiliations:** Medical University of South Carolina, Charleston, SC, USA; Medtronic, Inc, Mounds View, MN, USA; The Christ Hospital Health Network, Cincinnati, OH, USA

**Keywords:** Cardiac Resynchronization Therapy, Adaptive CRT, mortality, healthcare resource utilization, healthcare cost

## Abstract

**Objectives:** To assess the association between use of adaptive pacing on clinical and economic outcomes of CRT recipients in real-world analysis.

**Background:** The AdaptivCRT^TM^ algorithm was shown in prior subgroup analyses of prospective trials to achieve clinical benefits, but a large prospective trial showed nonsignificant changes in the endpoint of mortality or heart failure hospitalizations.

**Methods:** CRT-implanted patients from the Optum Clinformatics® database with ≥90 days of follow-up were included. Remote monitoring data was used to classify patients based on CRT setting – adaptive biventricular and left ventricular pacing (aCRT) vs. standard biventricular pacing (Standard CRT). Inverse probability of treatment weighting was used to adjust for baseline differences between groups. Mortality, 30-day readmissions, healthcare utilization, and payer and patient costs were evaluated post-implantation.

**Results:** This study included 2,412 aCRT and 1,638 Standard CRT patients (mean follow-up: 2.4 ± 1.4 years), with balanced baseline characteristics after adjustment. The aCRT group was associated with lower all-cause mortality (adjusted hazard ratio = 0.88 [95% confidence interval (CI):0.80, 0.96]), fewer all-cause 30-day readmissions (adjusted incidence rate ratio = 0.87 [CI:0.81, 0.94]), and fewer all-cause and HF-related inpatient, outpatient, and emergency department (ED) visits. The aCRT cohort was also associated with lower all-cause outpatient payer-paid amounts and lower all-cause and HF-related inpatient and ED patient-paid amounts.

**Conclusions:** In this retrospective analysis of a large real-world cohort, use of an adaptive CRT algorithm was associated with lower mortality, reduced healthcare resource utilization, and lower payer and patient costs.

## INTRODUCTION

In a subset of patients with heart failure (HF) with reduced ejection fraction, cardiac resynchronization therapy (CRT) improve exercise capacity, quality of life, and functional status, as well as reduces HF hospitalizations and mortality.[1–8] However, many patients are poorly responsive to this therapy.[9, 10] Consequently, one of the major strategies to maximize clinical and structural benefit of CRT is to optimize the timing of atrioventricular stimulation.[11] Although echocardiographic guidance for atrioventricular optimization is available, it is time-consuming, complex, costly, and lacks reproducibility and thus is seldom used in clinical practice.[12–15] Adaptive pacing algorithms (aCRT), which leverage a CRT programming strategy to preserve intrinsic atrio-ventricular (AV) conduction via the right bundle branch, mitigate the challenges with echocardiographic methods and allow for automated measurement of optimal pacing parameters.[16]

The Adaptive CRT study demonstrated non-inferiority of one aCRT, the AdaptivCRT^TM^ algorithm, compared with echocardiography-guided AV optimization in CRT patients;[17, 18] post-hoc analyses identified patients who may benefit further.[19] As such, the global, randomized AdaptResponse trial tested the hypothesis that use of the AdaptivCRT algorithm lowers the incidence of composite all-cause mortality or intervention for heart failure decompensation in patients with left bundle branch block (LBBB) and normal AV conduction in a clinical setting.[18, 20–23] While an 11% lower risk of composite all-cause mortality or acute heart failure intervention was observed in the aCRT arm, this difference did not reach statistical significance (HR=0.89, 95% CI: 0.78-1.01; p=0.077).

The AdaptResponse trial had lower-than-expected observed event rates, which may be a consequence of its narrow inclusion criteria, including only CRT-indicated patients with LBBB and normal AV conduction. By contrast, the Adaptive CRT study, which did observe significantly lower composite mortality or heart failure hospitalization in post-hoc analysis, enrolled a broader, CRT-indication population.[18] The lower-than expected event rates in both study arms may suggest that there was a greater than expected response to CRT in the included population.

With mixed evidence in disparate patient populations, limited evidence exists on the use of aCRT in a real-world population, as well as the implications for healthcare utilization and healthcare costs. Thus, this analysis aimed to evaluate the potential effect of aCRT on mortality, healthcare resource utilization (HRU), and healthcare costs leveraging a large, real-world, dataset derived from device registries and healthcare administrative claims.

## METHODS

### Study design

This was a retrospective, observational cohort study which was conducted to compare outcomes between patients with the AdaptivCRT algorithm (aCRT group) versus standard biventricular (BiV) pacing (Standard CRT group) (Figure 1). The choice of therapy setting was left to the clinicians caring for the patients. The index date was defined as the date associated with the first observed CRT implant procedure that occurred between June 1, 2013 (when the AdaptivCRT algorithm came into common use) and December 31, 2019. Patients with a recorded CRT implantation and confirmed patient linkage across the CIED device registry and Optum data sources (see “Data” section below), were at least 18 years old on the index date, had continuous insurance eligibility during the baseline period, and had at least 90 days of follow-up, were eligible for inclusion in this study. Patients with missing data on sex were excluded. The baseline period was defined as the 180-day interval prior to the index date. The follow-up period began on the day following the index date and continued until the end of the patient’s continuous eligibility in an insurance plan, end of data cut (December 31, 2020) or death. Treatment group assignment was based on the last observed CRT setting within 7 days post implant obtained from the Medtronic CareLink^®^ database. A patient was assigned into either the aCRT group when the observed CRT setting was “Adaptive BiV and LV” or the Standard CRT group when the observed CRT setting was “Nonadaptive CRT.” Patients whose CRT setting was “Adaptive BiV only” were excluded from the analysis.

**Figure 1.**
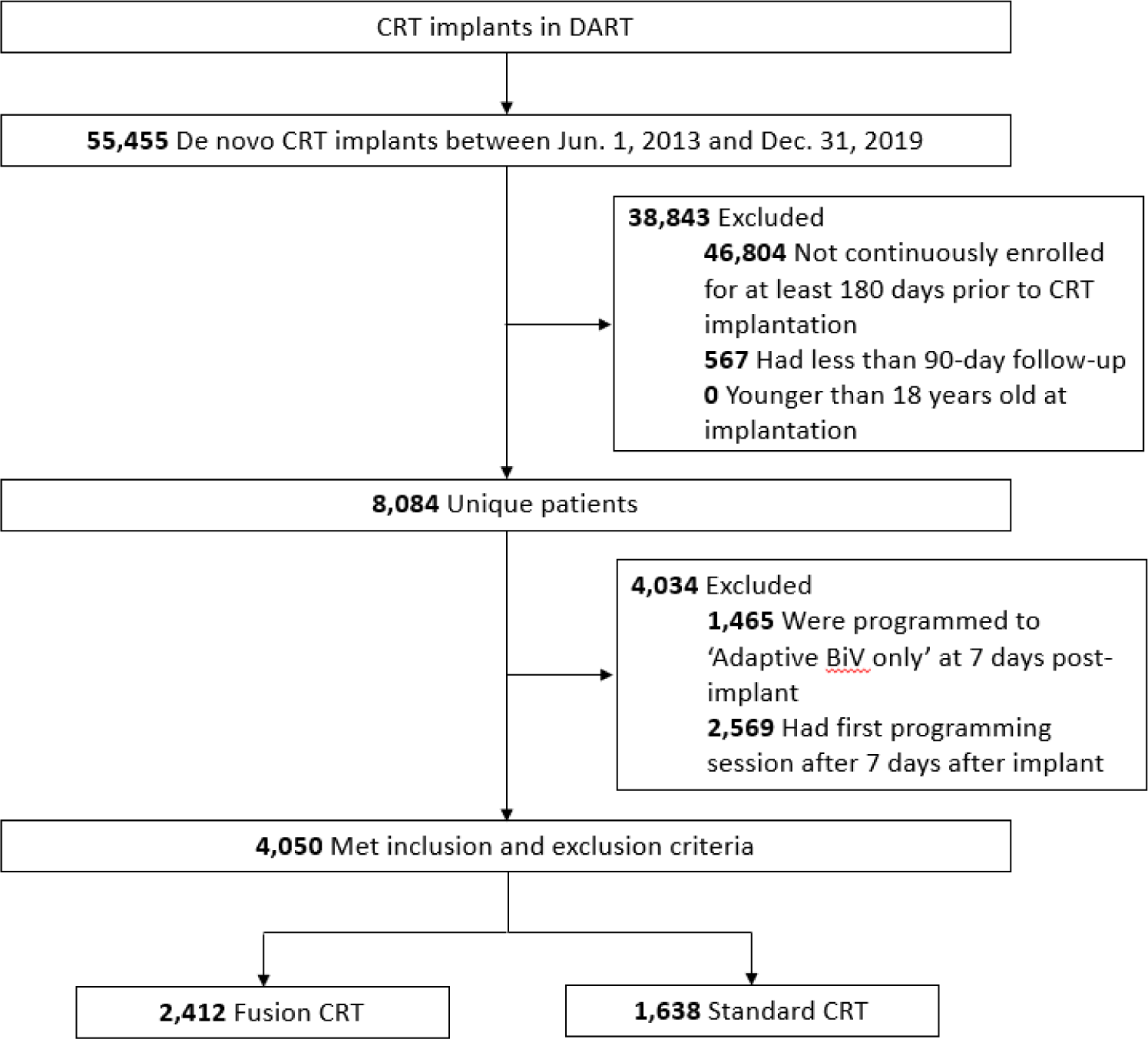
Flow chart of the study sample showing inclusion and exclusion of patients

### Data

Data from Optum’s de-identified Clinformatics^®^ Data Mart Database (CDM) were used. The database comprises four data sources linked at the patient level, including commercial and Medicare Advantage health plan data for approximately 61 million unique members from all US census regions, Social Security Death Index data, the Medtronic CareLink^®^ remote monitoring data warehouse, and the Medtronic Device and Registrant Tracking system. The CDM includes enrollment records, medical claims, and prescription claims. The CDM is statistically de-identified under the Expert Determination method consistent with the Health Information Insurance Portability and Accountability Act and managed according to Optum^®^ customer data use agreements. The CDM administrative claims submitted for payment by providers and pharmacies are verified, adjudicated, adjusted, and de-identified prior to inclusion. This study was deemed exempt from review by the Institutional Review Board at Medtronic because this study used only de-identified administrative health claims and did not meet the definition of human subject research.

### Patient characteristics

Patient demographic characteristics were collected on the index date, including age, sex, geographic region, insurance type, and year of index date. Baseline comorbidities (see Table 1) were identified using ICD-9-CM and ICD-10-CM diagnoses codes in medical claims. The Charlson comorbidity index was used to measure the severity of comorbid conditions.[21] Use of medications (i.e., beta-blockers, loop diuretics, angiotensin converting enzyme inhibitors, angiotensin receptor blockers) during the baseline period were identified using Healthcare Common Procedure Coding System codes in medical claims and national drug codes in prescription claims. In a sensitivity analysis, patients who had evidence of pacemaker and implantable cardioverter defibrillator (ICD) implants during the baseline period were excluded. Details of the diagnosis, procedure, and drug codes used are provided in Supplementary Tables A1-A4.

**Table 1.**
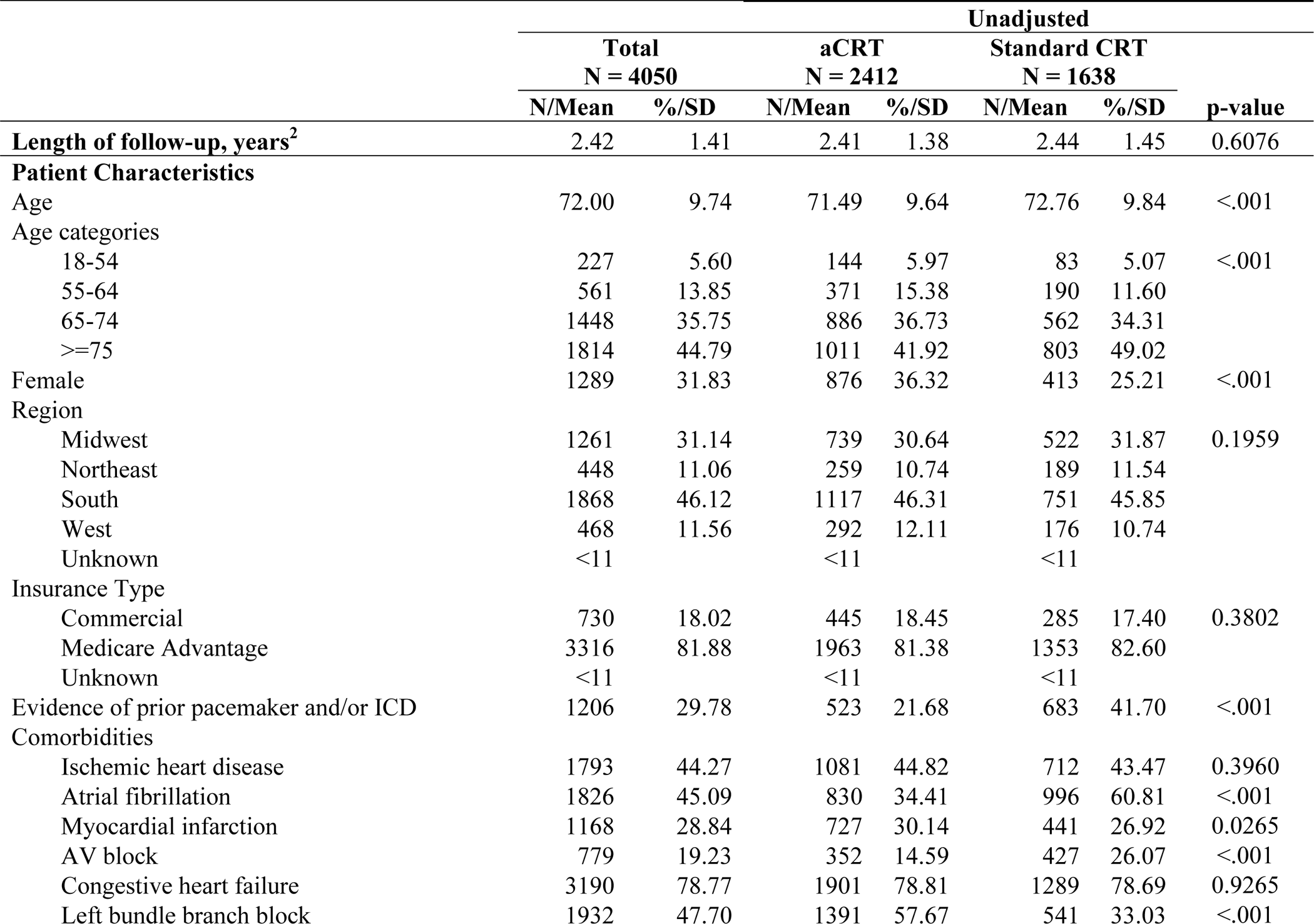
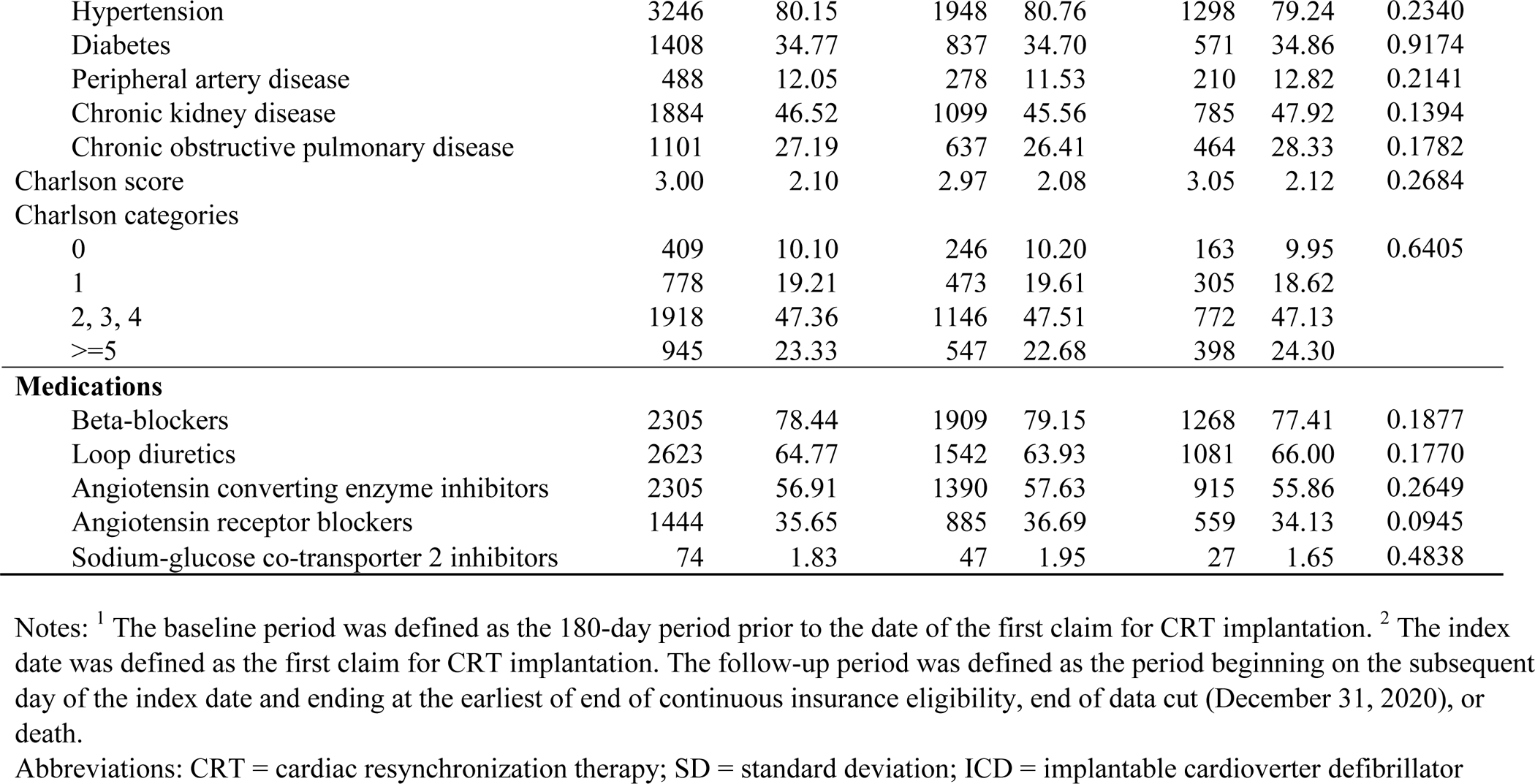
Patient characteristics during the baseline period^1^.

|  | Unadjusted |  |  |  |  |  |  |
| --- | --- | --- | --- | --- | --- | --- | --- |
|  | Total<br>N = 4050 |  | aCRT<br>N = 2412 |  | Standard CRT<br>N = 1638 |  | p-value |
|  | N/Mean | %/SD | N/Mean | %/SD | N/Mean | %/SD |  |
| **Length of follow-up, years<sup>2</sup>** | 2.42 | 1.41 | 2.41 | 1.38 | 2.44 | 1.45 | 0.6076 |
| **Patient Characteristics** |  |  |  |  |  |  |  |
| Age | 72.00 | 9.74 | 71.49 | 9.64 | 72.76 | 9.84 | <.001 |
| Age categories |  |  |  |  |  |  |  |
| 18-54 | 227 | 5.60 | 144 | 5.97 | 83 | 5.07 | <.001 |
| 55-64 | 561 | 13.85 | 371 | 15.38 | 190 | 11.60 |  |
| 65-74 | 1448 | 35.75 | 886 | 36.73 | 562 | 34.31 |  |
| ≥75 | 1814 | 44.79 | 1011 | 41.92 | 803 | 49.02 |  |
| Female | 1289 | 31.83 | 876 | 36.32 | 413 | 25.21 | <.001 |
| Region |  |  |  |  |  |  |  |
| Midwest | 1261 | 31.14 | 739 | 30.64 | 522 | 31.87 | 0.1959 |
| Northeast | 448 | 11.06 | 259 | 10.74 | 189 | 11.54 |  |
| South | 1868 | 46.12 | 1117 | 46.31 | 751 | 45.85 |  |
| West | 468 | 11.56 | 292 | 12.11 | 176 | 10.74 |  |
| Unknown | <11 |  | <11 |  | <11 |  |  |
| Insurance Type |  |  |  |  |  |  |  |
| Commercial | 730 | 18.02 | 445 | 18.45 | 285 | 17.40 | 0.3802 |
| Medicare Advantage | 3316 | 81.88 | 1963 | 81.38 | 1353 | 82.60 |  |
| Unknown | <11 |  | <11 |  | <11 |  |  |
| Evidence of prior pacemaker and/or ICD | 1206 | 29.78 | 523 | 21.68 | 683 | 41.70 | <.001 |
| Comorbidities |  |  |  |  |  |  |  |
| Ischemic heart disease | 1793 | 44.27 | 1081 | 44.82 | 712 | 43.47 | 0.3960 |
| Atrial fibrillation | 1826 | 45.09 | 830 | 34.41 | 996 | 60.81 | <.001 |
| Myocardial infarction | 1168 | 28.84 | 727 | 30.14 | 441 | 26.92 | 0.0265 |
| AV block | 779 | 19.23 | 352 | 14.59 | 427 | 26.07 | <.001 |
| Congestive heart failure | 3190 | 78.77 | 1901 | 78.81 | 1289 | 78.69 | 0.9265 |
| Left bundle branch block | 1932 | 47.70 | 1391 | 57.67 | 541 | 33.03 | <.001 |
| Hypertension | 3246 | 80.15 | 1948 | 80.76 | 1298 | 79.24 | 0.2340 |
| Diabetes | 1408 | 34.77 | 837 | 34.70 | 571 | 34.86 | 0.9174 |
| Peripheral artery disease | 488 | 12.05 | 278 | 11.53 | 210 | 12.82 | 0.2141 |
| Chronic kidney disease | 1884 | 46.52 | 1099 | 45.56 | 785 | 47.92 | 0.1394 |
| Chronic obstructive pulmonary disease | 1101 | 27.19 | 637 | 26.41 | 464 | 28.33 | 0.1782 |
| Charlson score | 3.00 | 2.10 | 2.97 | 2.08 | 3.05 | 2.12 | 0.2684 |
| Charlson categories |  |  |  |  |  |  |  |
| 0 | 409 | 10.10 | 246 | 10.20 | 163 | 9.95 | 0.6405 |
| 1 | 778 | 19.21 | 473 | 19.61 | 305 | 18.62 |  |
| 2, 3, 4 | 1918 | 47.36 | 1146 | 47.51 | 772 | 47.13 |  |
| >=5 | 945 | 23.33 | 547 | 22.68 | 398 | 24.30 |  |
| **Medications** |  |  |  |  |  |  |  |
| Beta-blockers | 2305 | 78.44 | 1909 | 79.15 | 1268 | 77.41 | 0.1877 |
| Loop diuretics | 2623 | 64.77 | 1542 | 63.93 | 1081 | 66.00 | 0.1770 |
| Angiotensin converting enzyme inhibitors | 2305 | 56.91 | 1390 | 57.63 | 915 | 55.86 | 0.2649 |
| Angiotensin receptor blockers | 1444 | 35.65 | 885 | 36.69 | 559 | 34.13 | 0.0945 |
| Sodium-glucose co-transporter 2 inhibitors | 74 | 1.83 | 47 | 1.95 | 27 | 1.65 | 0.4838 |

Notes: <sup>1</sup> The baseline period was defined as the 180-day period prior to the date of the first claim for CRT implantation. <sup>2</sup> The index date was defined as the first claim for CRT implantation. The follow-up period was defined as the period beginning on the subsequent day of the index date and ending at the earliest of end of continuous insurance eligibility, end of data cut (December 31, 2020), or death.
Abbreviations: CRT = cardiac resynchronization therapy; SD = standard deviation; ICD = implantable cardioverter defibrillator

### Outcomes

The outcomes of this study were assessed during the follow-up period and included all-cause mortality, all-cause and HF-related hospitalizations, 30-day readmissions, HRU and payer and patient healthcare costs. An all-cause 30-day readmission was defined as an inpatient visit with an admission date within 30 days of a previous discharge from an inpatient visit that itself was not a readmission. A HF-related 30-day readmission was defined as an inpatient visit within 30 days of discharge from a HF-related index inpatient visit.[21]

HRU and healthcare costs were categorized by place of service (inpatient, outpatient, emergency department, and other). A hierarchy was used to categorize place of service for patients with multiple visits on the same day, prioritizing inpatient, then emergency department, then outpatient, then other settings. Emergency department visits that resulted in inpatient admissions were categorized as inpatient visits and did not count as a separate encounter. The “other” category includes claims related to laboratory, durable medical equipment, diagnostic tests, and other professional services. Annual payer paid amounts and patient paid amounts were estimated using standardized medical costs computed from Optum^®^, coinsurance, copay, and deductible amounts. All costs were adjusted to 2020 USD using the Medical Care component of the US Census Bureau Consumer Price Index.

### Statistical analysis

Baseline characteristics were compared between the aCRT and Standard CRT groups using t-tests (normally distributed continuous variables), Wilcoxon-Mann-Whitney non-parametric tests (non-normally distributed continuous variables), or Chi-squire tests (categorical variables). HRU and costs were summarized in terms of annualized amounts per patient to account for varying lengths of follow-up across patients.

To mitigate confounding, inverse probability of treatment weighting (IPTW) was used to adjust for differences in baseline characteristics between groups. We first estimated propensity scores (PS) using a logistic regression model in which treatment status was regressed on the following baseline characteristics: age, sex, geographic region, type of insurance plan, year of index date, comorbidities, Charlson comorbidity index, medication use, and evidence of pacemaker and ICD implants prior to their index CRT implant. Then, weights were calculated based on the inverse of the PS (IPTW = 1/PS for patients with aCRT; IPTW = 1/(1-PS) for patients with Standard CRT).[24] The balance of baseline characteristics between the two groups was compared using standardized mean differences (SMDs). A SMD lower than 10% was considered to represent no consequential imbalance between the two groups.[25]

Unadjusted and IPTW-adjusted comparisons of mortality were assessed using a hazard ratio (HR) estimated from Cox proportional hazards models. All-cause and HF-related readmissions and HRU were compared between groups through incidence rate ratios (IRR) estimated using multivariable Poisson regression models. Finally, healthcare costs were compared between groups through cost ratios using multivariable Tweedie regression models with patient-level annual costs as the dependent variable. Robust variance estimators were used to generate 95% confidence intervals (CI) and p-values. A two-tailed p < 0.05 was considered statistically significant. The analyses were performed using SAS 9.4 (SAS Institute, Cary, North Carolina).

## RESULTS

### Baseline Characteristics

Baseline characteristics for the full cohort as well as for each subgroup are presented in Table 1. The unadjusted parameters as well as their corresponding SMDs after the IPTW adjustment are shown. The included cohort was typical of CRT populations with approximately 80% being 65 years or older and one third female sex. Less than half (44%) of the population had ischemic heart disease, and 48% had a LBBB on the baseline electrocardiogram. As expected, hypertension, diabetes, atrial fibrillation, and chronic kidney disease were all prevalent co-morbidities in this population. The final cohort consisted of 2,412 patients with aCRT and 1,638 with Standard CRT. The mean follow-up duration was 2.4 years across all patients and was not significantly different between the comparator groups (p=0.61).

There were some important differences between the groups. Specifically, patients with aCRT (i.e., AdaptivCRT algorithm) were younger, more likely to be female, less likely to have evidence of a previous pacemaker or ICD (p<.001), and more likely to have evidence of LBBB (p<.001). With respect to comorbidities, patients with aCRT were more likely to have previous myocardial infarction (p<.05) and less likely to have evidence of atrial fibrillation and atrioventricular block (p<.001). The small resulting differences after adjustment (SMDs after IPTW) shows that this methodology appropriately balanced baseline characteristics (Supplementary Table A5).

### Clinical Outcomes

The adjusted and unadjusted HR from the comparison of mortality risk between the aCRT and Standard CRT groups are shown in Figure 2. There was a total of 483 (20.02%) and 471 (28.75%) deaths for any cause in the aCRT and Standard CRT groups, respectively. aCRT was associated with a 12% reduction in risk for all-cause mortality (adjusted HR [95% CI] = 0.88 [0.78, 0.96], p<.001). Figure 3 shows the adjusted survival curves for all-cause mortality, which yielded 1-, 3-, and 5-year survival rates of 0.96 (CI: 0.95, 0.96), 0.82 (CI: 0.80, 0.83), and 0.61 (CI: 0.57, 0.65), respectively, in the aCRT group, and 0.94 (CI: 0.93, 0.95), 0.75 (CI: 0.72, 0.77), and 0.54 (0.50, 0.58), respectively in the standard CRT group.

**Figure 2.**
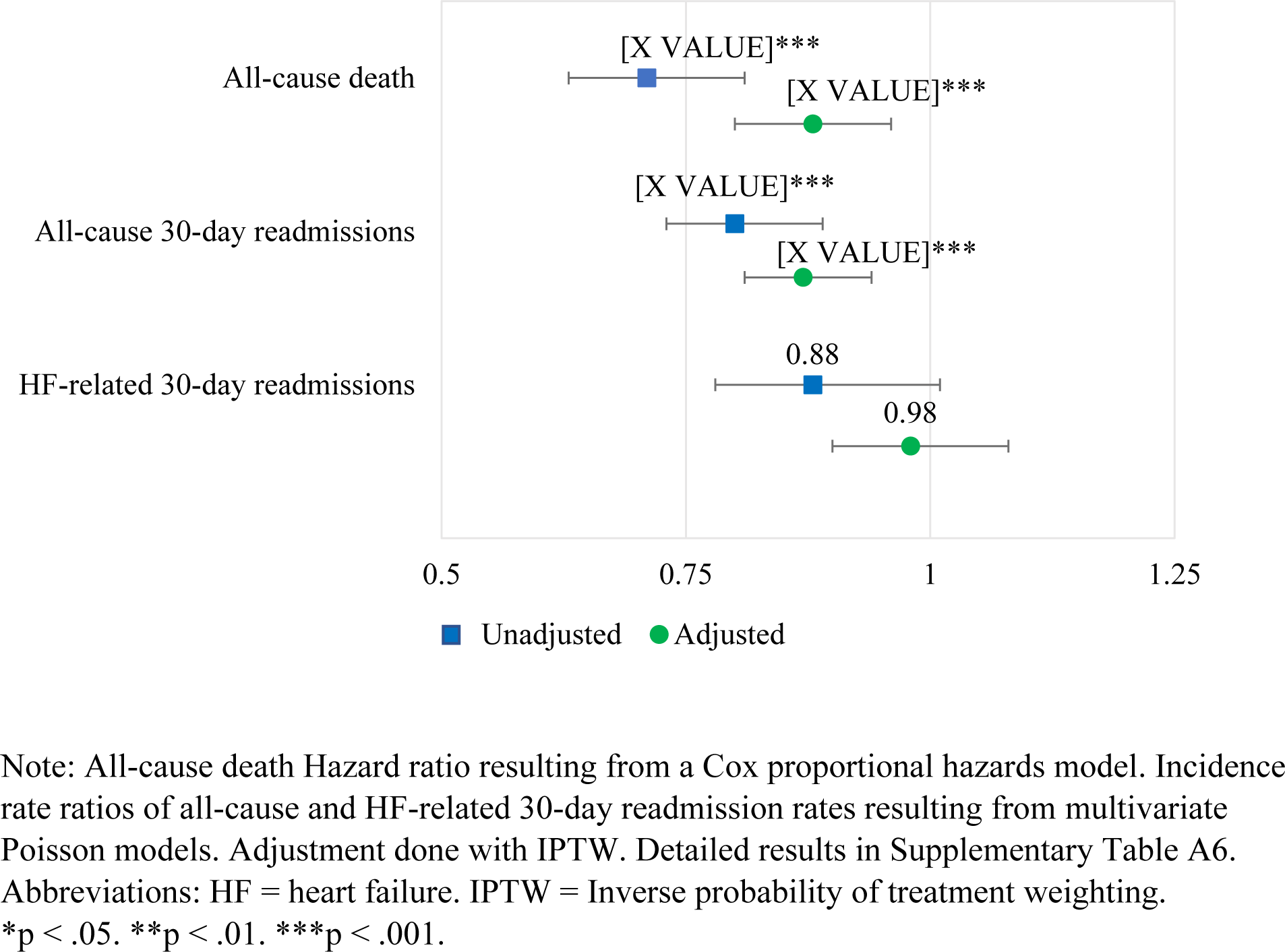
Hazard Ratios and Incidence Rate Ratios of aCRT vs. Standard CRT.

**Figure 3.**
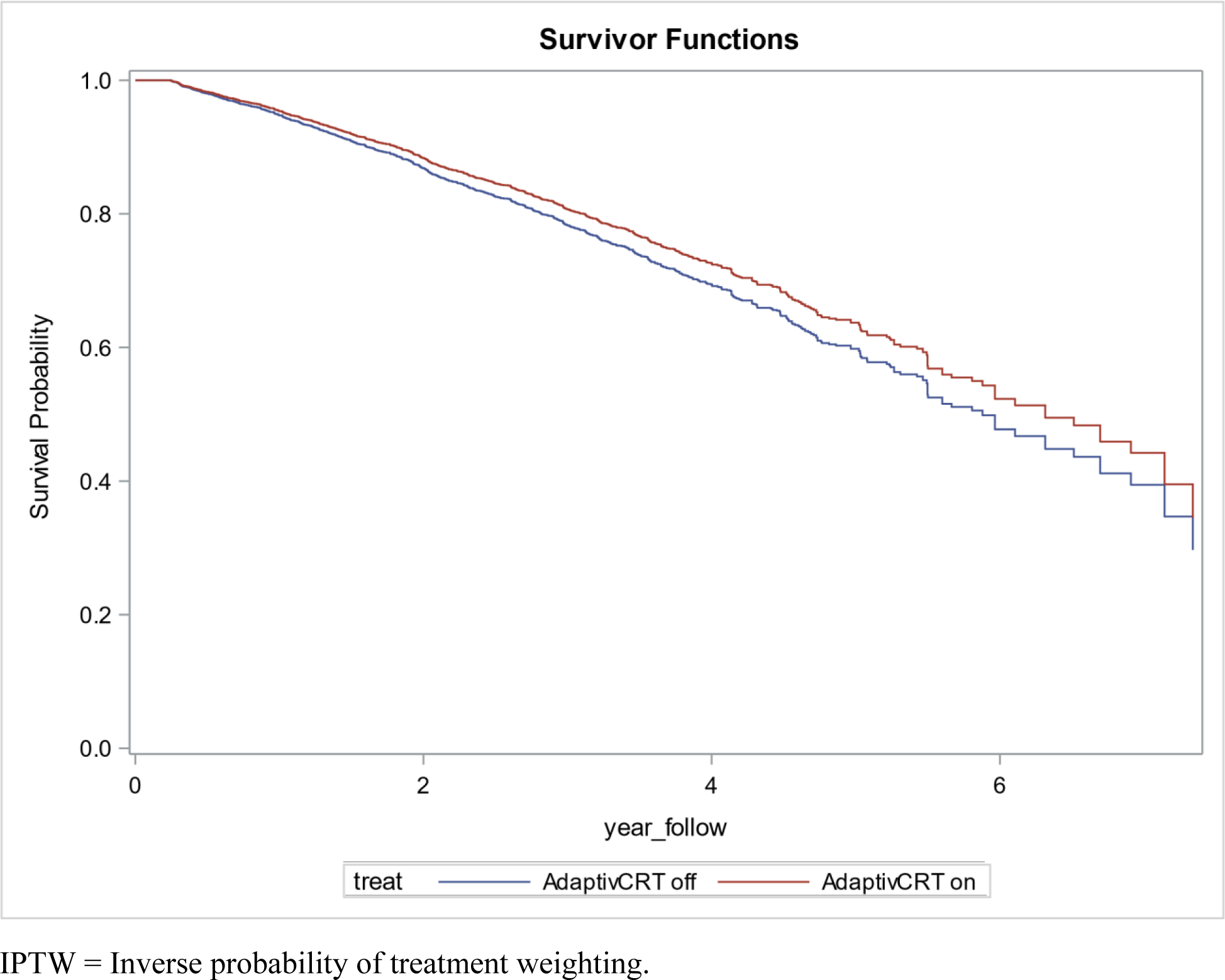
IPTW-Adjusted survivor functions from Cox Proportional Hazard model of all-cause mortality.

The annual all-cause and HF-related 30-day readmission rates in the aCRT group were 0.21 per patient-year and 0.15 per patient-year, respectively, compared to 0.28 per patient-year and 0.17 per patient-year in the Standard CRT group. Figure 2 shows the adjusted and unadjusted IRR of the 30-day readmission rates from Poisson models. aCRT was associated with a 13% lower all-cause 30-day readmission rate compared to Standard CRT (adjusted incidence rate ratio (aIRR) [95% CI] = 0.87 [0.81, 0.94], p<.001). HF-related 30-day readmissions were not significantly different between the two groups (aIRR = 0.98 [0.90,1.08], p=0.74).

### Health Care Utilization

With respect to healthcare utilization, aCRT was associated with lower all-cause inpatient (aIRR = 0.94 [0.90, 0.98], p<.01), outpatient (aIRR = 0.85 [0.80, 0.91], p<.001), and emergency department (aIRR = 0.78 [0.68, 0.89], p<.001) visits. These results are summarized in Figure 4. Similarly, aCRT was associated with lower HF-related inpatient (aIRR = 0.95 [0.91, 0.99], p<.05), outpatient (aIRR = 0.87 [0.81, 0.95], p<.001), and emergency department (aIRR = 0.72 [0.59, 0.88], p<.01) visits.

**Figure 4.**
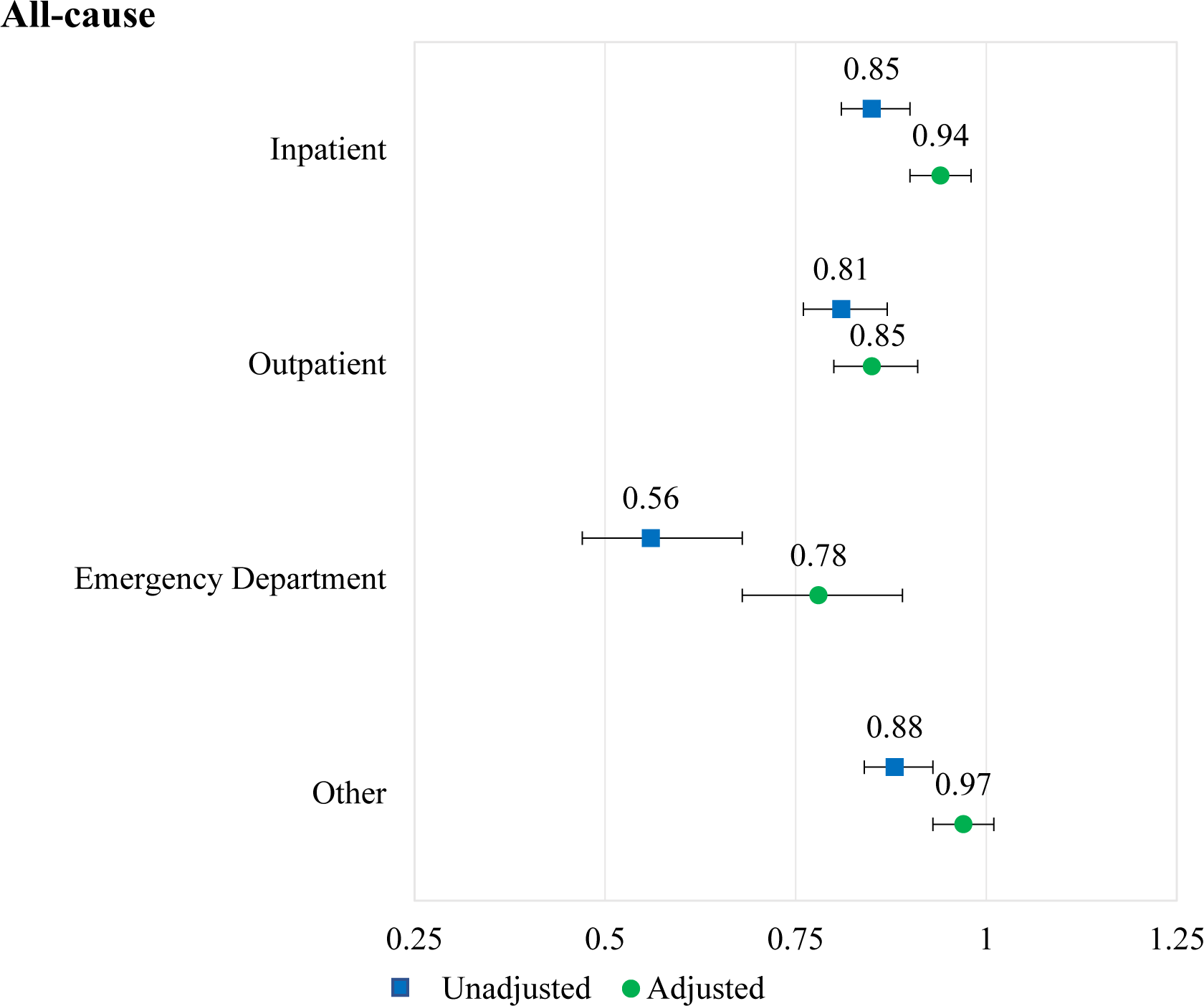
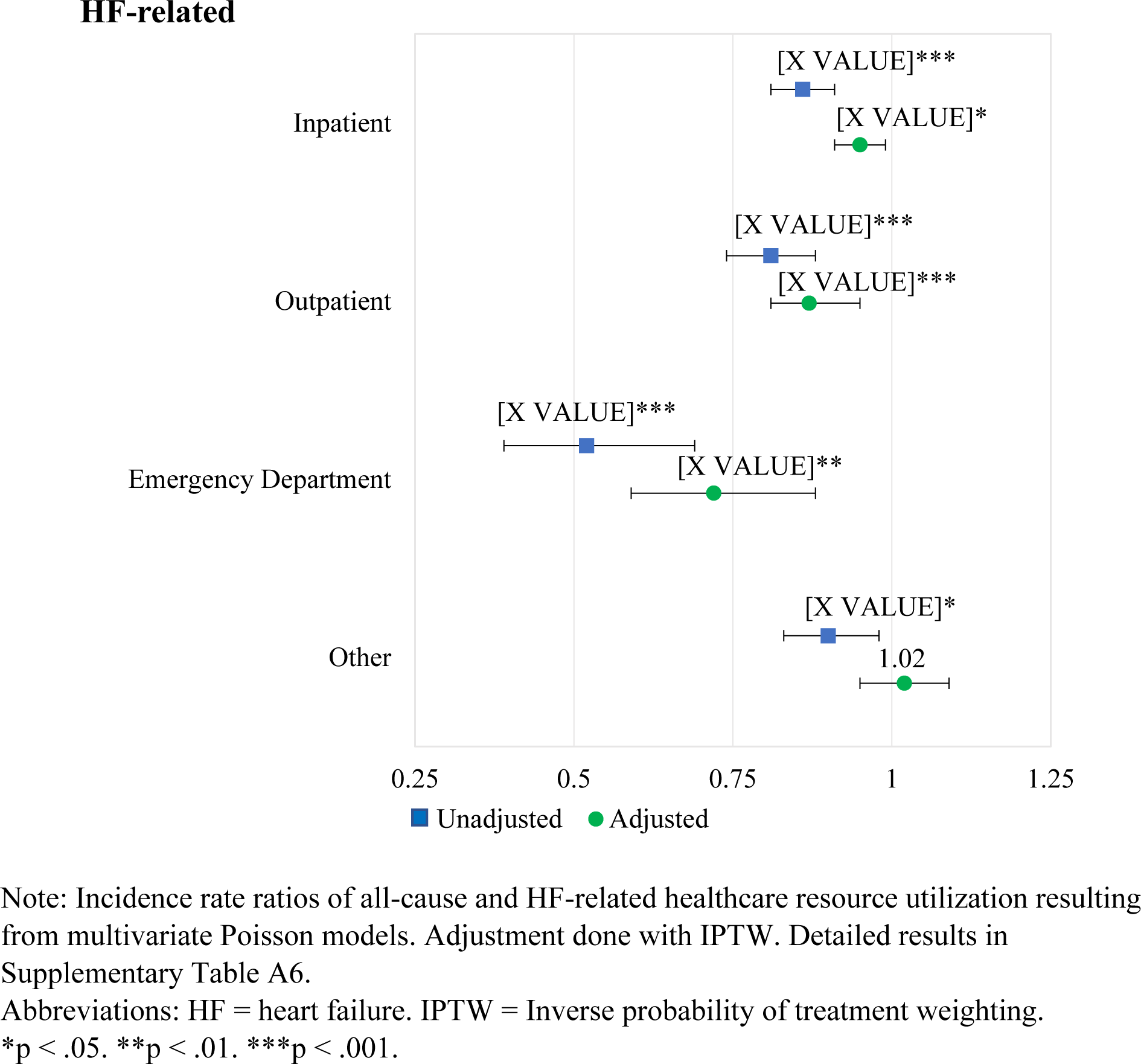
Health Resource Utilization Incidence Rate Ratios of aCRT vs. Standard CRT.

Adjusted mean annual all-cause per-patient payer-paid amounts ranged from $19,820 for inpatient care to $138 for emergency care in patients with aCRT and from $22,674 for inpatient care to $296 for emergency care in patients with Standard CRT. The mean HF-related annual payer costs for aCRT and Standard CRT ranged from $17,474 and $19,359, respectively, for inpatient care to $60 and $100, respectively, for emergency care. Mean (standard deviation) annual all-cause outpatient payer costs were significantly lower for aCRT compared to Standard CRT ($10,505 [$26,969] vs. $11,748 [$34,842], adjusted Cost Ratio [95% CI] = 0.92 [0.85, 0.99], p<.05) (Figure 5a). aCRT was also associated with lower annual inpatient and emergency department patient costs relative to Standard CRT (p<.05) (Figure 5b). No other statistically significant differences were observed across other categories of payer or patient healthcare costs following adjustment.

**Figure 5a.**
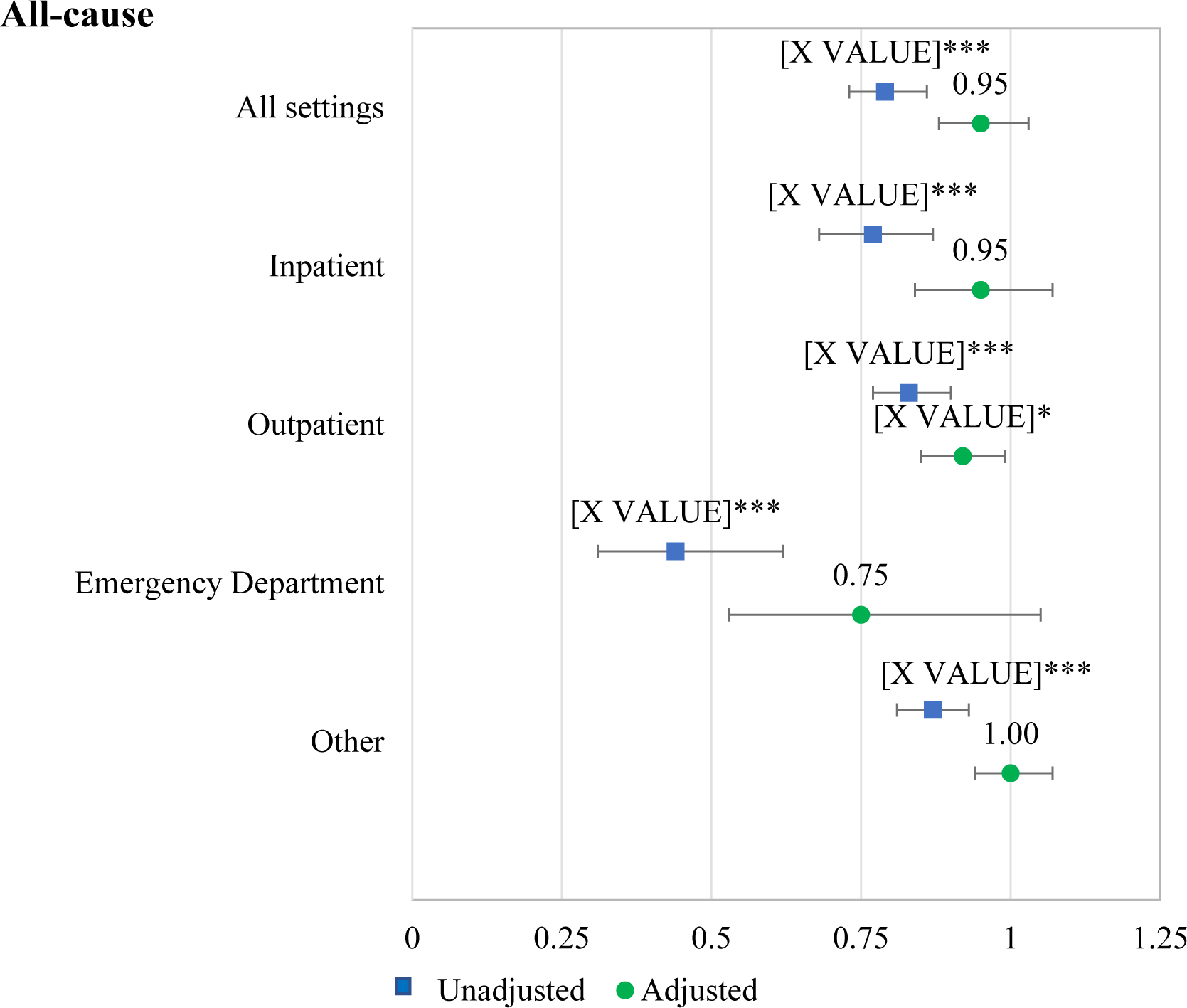
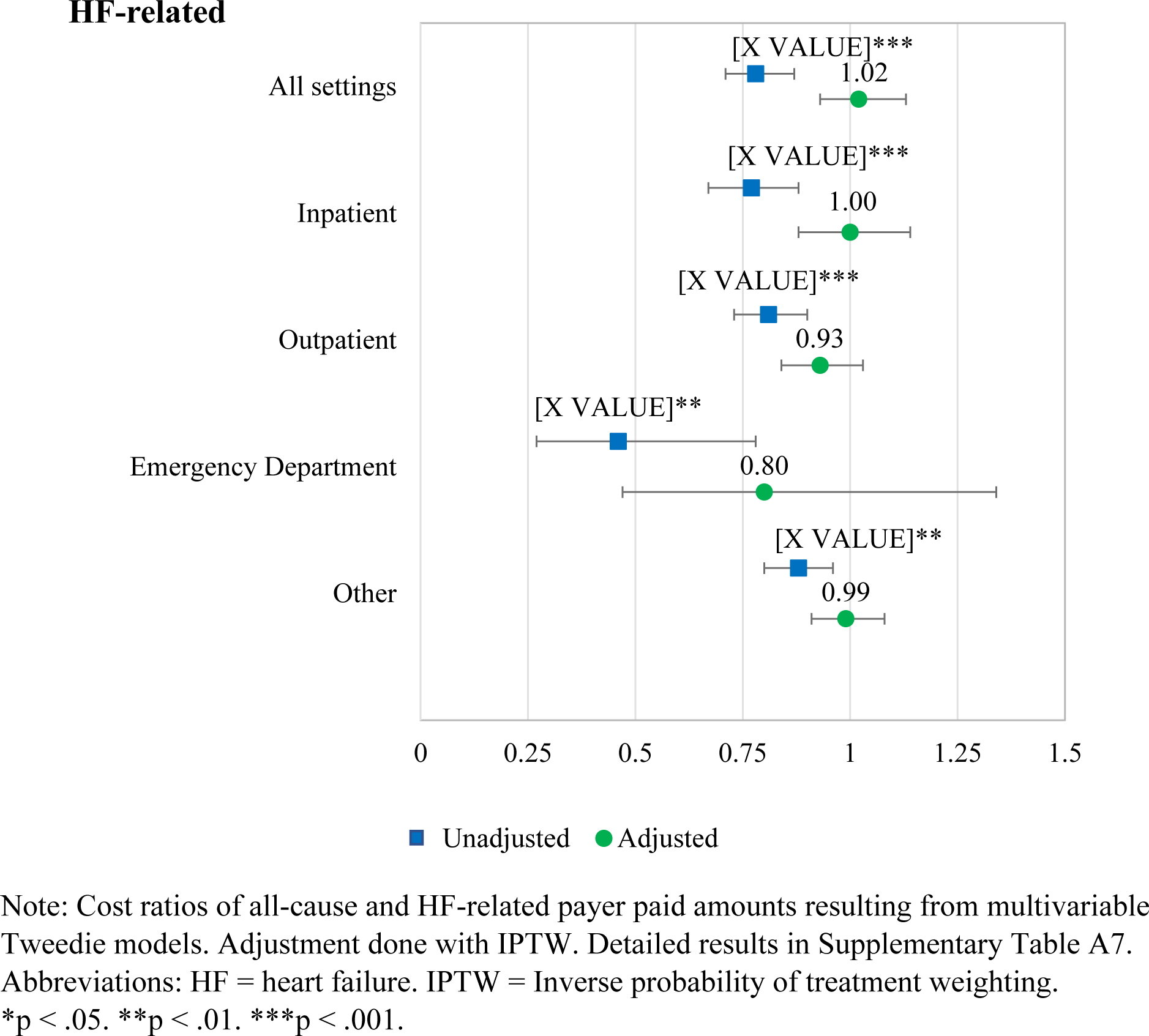
Payer Cost Ratios of aCRT vs. Standard CRT

**Figure 5b.**
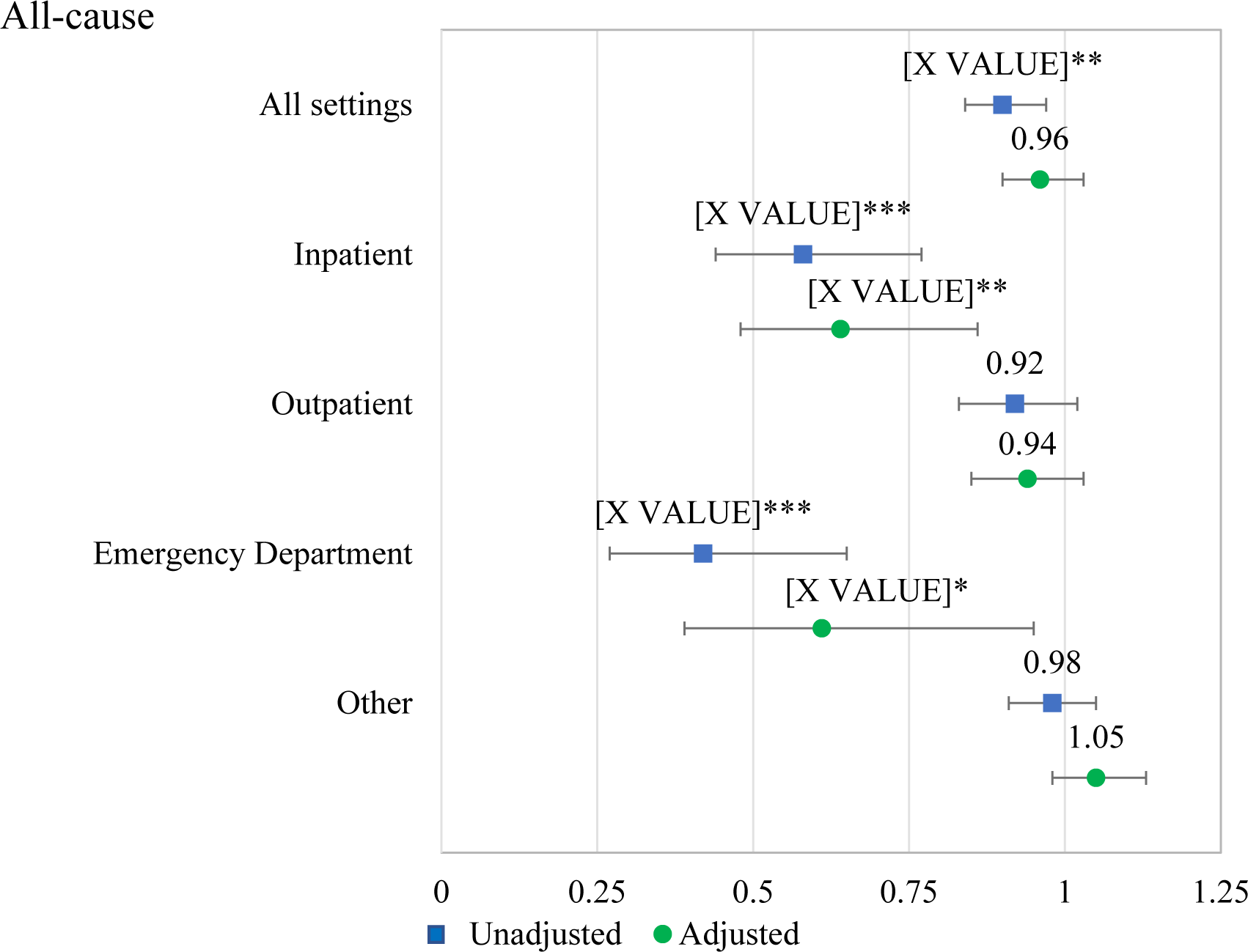
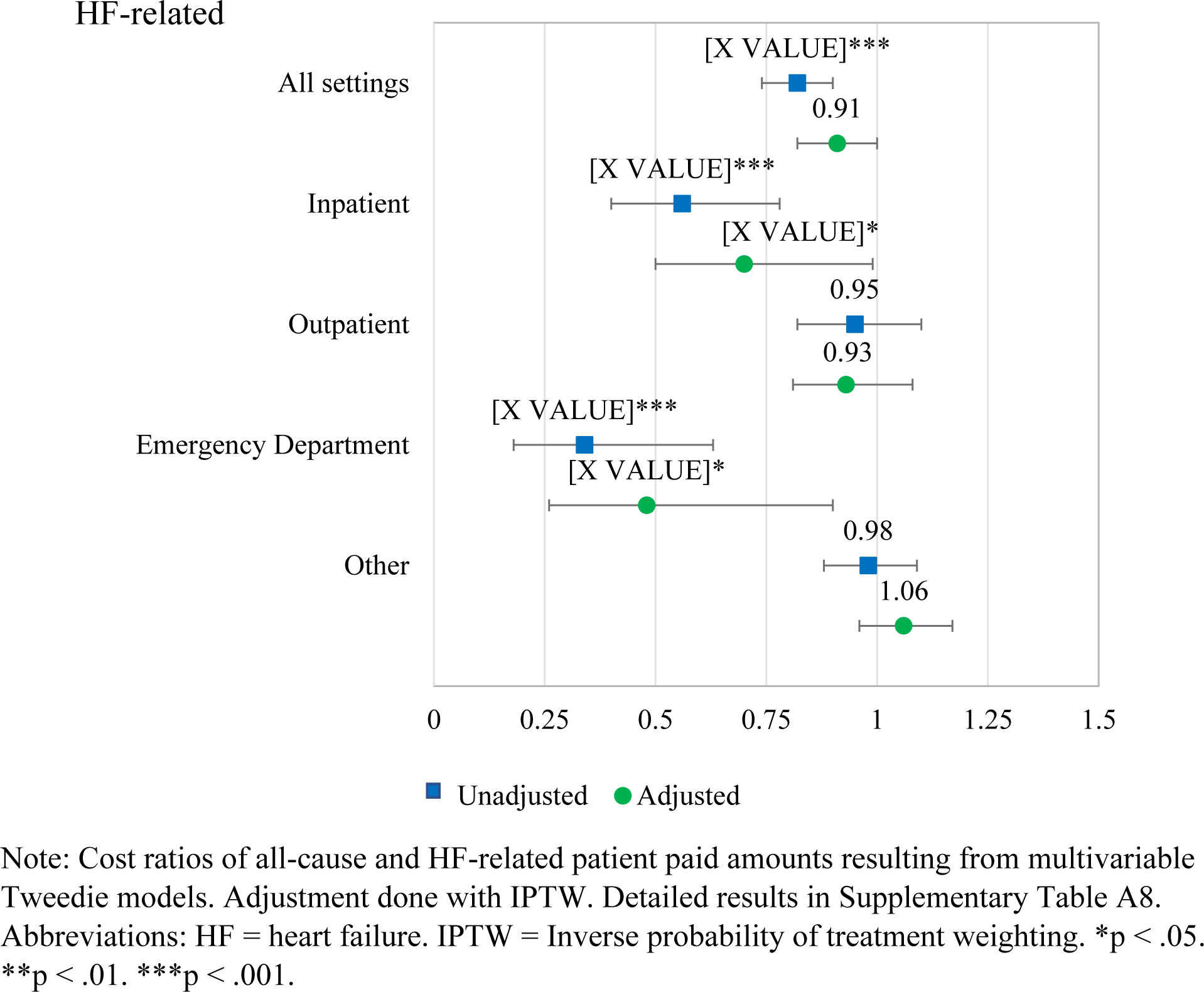
Patient Cost Ratios of aCRT vs. Standard CRT

## DISCUSSION

The present study leverages a large administrative database to assess the potential real-world impact of an adaptive CRT pacing algorithm on clinical outcomes, HRU, and costs in clinical practice. This is the first real-world claims-based analysis of a pacing algorithm designed to provide fusion pacing from the left ventricle. There are 5 important findings from the study: The aCRT algorithm (Adaptive BiV and LV pacing) was associated with **1)** reduced all-cause mortality, **2)** reduced all-cause 30-day readmissions, **3)** lower HRU in the inpatient, outpatient, and emergency department settings, **4)** lower payer costs (for all-cause outpatient and emergency department visits) and, **5)** lower patient costs (for all-cause and HF-related inpatient and emergency department visits).

Our findings contribute to a growing body of evidence on the health and economic effects of aCRT. Early evidence was largely positive with the randomized Adaptive CRT trial demonstrating non-inferior clinical outcomes in aCRT compared to echocardiography-optimized patients,[18] as well as a subsequent post-hoc analysis by Starling et al. which showed a relative risk reduction in 30-day readmissions (59% for HF-related and 46% for all-cause readmissions) with aCRT.[22] A subsequent post-market registry revealed a 29% relative risk reduction in mortality with aCRT when compared to standard BiV pacing patients.[26] Other studies have found mortality benefits in HF patients with aCRT compared to traditional CRT.[20, 27]

By contrast, in the AdaptResponse trial, aCRT, compared with conventional CRT, did not significantly reduce all-cause death or intervention for heart failure decompensation in a select cohort of HF patients with LBBB and intact AV conduction. However, AdaptResponse differed from the present analysis with respect to both the population included and outcomes examined. AdaptResponse enrolled a population that was, on average, 7 years younger and all patients had LBBB compared to 48% in the present analysis – although, the low LBBB percentage in this study may have been partially due to under diagnosis and under-documentation in administrative claims data. The neutral findings likely were a result in part to a lower-than-expected rate of primary outcome events within this highly selected CRT population. Specifically, the 5-year Kaplan-Meier estimates for the risk of death in AdaptResponse were 15.6% and 17.4% in the aCRT and standard CRT groups, respectively. By contrast, in the present analysis, the risk of death at 5 years was 39% in the aCRT group and 46% of the Standard CRT group. Notably, while the relative reduction in mortality was nearly identical (12% reduction in both studies) and sample sizes were similar (3,617 in AdaptResponse vs. 4,050 the present analysis), this reduction was highly significant in the present analysis (p<0.001) but did not achieve statistical significance in AdaptResponse (p=0.12), illustrating the increase in statistical power with the higher event rates in the current analysis. In contrast to most randomized trials, the current real-world clinical practice cohort was comprised of older patients with more atrial fibrillation and less LBBB. Nevertheless, this analysis also observed a 12% lower risk of all-cause mortality and 13% lower rate of all-cause 30-day readmission in the aCRT group compared to the Standard CRT group. Contrary to the findings of the Adaptive CRT trial, yet consistent with the AdaptResponse trial, the current analysis did not find a reduction in HF-related 30-day readmissions. This difference may be attributed to a combination of differences in populations studied, as well as the controlled nature of the prior study with specific adaptive pacing programming requirements and event reporting and adjudication.

The present study observed that aCRT was associated with significant reductions in both all-cause and HF-related HRU across inpatient, outpatient and emergency department settings.[26, 28] The clinical benefits associated with aCRT may also confer an economic benefit. One previous economic study projected a significant cost offset in the United States associated with aCRT.[29] A Japanese-fee schedule based cost minimization analysis demonstrated that aCRT was associated with significant cost offsets compared to traditional CRT.[30, 31] In the present study, aCRT was associated with lower payer-paid amounts for all-cause outpatient care and patient cost reductions in outpatient and emergency visits. While this is promising, a study designed to provide additional economic data, such as a subsequent cost analysis of the AdaptResponse trial, is needed to further assess the cost effectiveness and economic value of aCRT.

This study should be interpreted in light of several limitations. First, the results of this study refer to an intent-to-treat analysis, as adaptive pacing mode could change during the follow-up period. We applied a 7-day run-in period to assign patients to either the aCRT or Standard CRT groups. During the first 7 days post-implant, only 15 (0.4%) patients’ devices were reprogrammed. Second, as a retrospective analysis based on administrative claims data, aCRT status was not randomly assigned, and the outcomes are not adjudicated to aCRT status. However, the IPTW adjustment corrects for observed confounders and reduces the biases introduced by non-random treatment assignment. Lastly, the claims data are not representative of all HF populations in the US, so the results of this analysis cannot be extrapolated to the whole HF population in the US.

## CONCLUSIONS

Using real-world data from commercial and Medicare Advantage patients implanted with CRT devices, our analysis found patients with an adaptive CRT algorithm to have a significantly lower all-cause mortality rate, lower all-cause 30-day readmission rates, lower all-cause and HF-related HRU across multiple care settings, and lower healthcare costs than similar patients with traditional CRT. There was no significant difference in HF-related 30-day readmission rates with aCRT.

## Data Availability

The Optum Clinformatics database is available for purchase through Optum. However, the Optum Clinformatics database linked with Medtronic device registry data is only available to Medtronic personnel.

## Acknowledgements

The authors thank Randy Crossland, PhD, Dedra Fagan, PhD, Xiaoxiao Lu, PhD, and Joshua Ikuemonisan, MD of Medtronic, Inc. for assistance in the preparation of this manuscript.

## ABBREVIATION LIST

aCRT: Adaptive CRT
aIRR: Adjusted Incidence Rate Ratio
AV: Atrio-ventricular
BiV: Biventricular
CDM: Clinformatics® Data Mart
CI: Confidence intervals
CRT: Cardiac resynchronization therapy
HF: Heart failure
HR: Hazard ratio
HRU: Healthcare resource utilization
ICD: Implantable cardioverter defibrillator
ICD-9-CM: The International Classification of Disease, Ninth Revision, Clinical Modification
ICD-10-CM: The International Classification of Disease, Tenth Revision, Clinical Modification
IPTW: Inverse probability of treatment weighting
IRR: Incidence rate ratio
LBBB: Left bundle branch block
LV: Left ventricular
PS: Propensity score
SMD: Standardized mean difference

